# Why is implementing remote monitoring in virtual wards (Hospital at Home) for people living with frailty so hard? Qualitative interview study

**DOI:** 10.1101/2024.10.02.24314760

**Authors:** Rosie Lindsay, Paige Cunnington, Mary Dixon-Woods

## Abstract

**Background:** There is relatively low uptake of remote monitoring on frailty virtual wards compared to virtual wards caring for people with other medical conditions. However, reasons for low uptake are poorly understood.

**Objectives:** To explore the views and experiences of frailty virtual wards stakeholders involved in implementing remote monitoring.

**Methods:** We conducted qualitative interviews with 42 stakeholders involved in delivery, design or evaluation of frailty virtual wards based in the United Kingdom (UK). Analysis was based on the constant comparative method.

**Results:** Many participants perceived that remote monitoring was only useful for a small sub-group of patients with frailty for a range of medical, practical and social reasons. Remote monitoring required new ways of working from patients, staff and carers. The nature of this work was not always sufficiently well understood, designed, or supported. Procurement practices were also seen to be mis-aligned with service needs, resulting in provision of equipment that was not fit for purpose. A further challenge in implementing remote monitoring in frailty virtual wards lay in tensions between national-level standardisation and enabling local flexibility.

**Conclusions:** Implementing remote monitoring in frailty virtual wards is challenged by lack of consensus on its suitability for this population, the extent and nature of change in clinical practices and work systems design required, and issues relating to equipment and standardisation. More co-design effort is needed to inform decision-making on remote monitoring for those with frailty.

## Introduction

Recent years have seen rapid growth worldwide in efforts to divert patients from hospital admission [1]. One major driver for this in the English National Health Service (NHS) is limited capacity, evident in very high occupancy rates (e.g. 94.9% in English hospitals in March 2024 [2] compared with a recommended threshold of 85% [3]). The urge to avert or reduce hospitalisation has been given further impetus by growing recognition of the negative consequences of hospital stays, including risks of healthcare-associated infections [4], people being placed in unsafe or undignified conditions owing to lack of beds [5], and deconditioning, particularly for older people and those living with frailty [6].

Among the prominent alternatives to hospitalisation are interventions such as Hospital at Home and virtual wards. The two terms are often used interchangeably. The various definitions have in common that they refer to care that is normally provided in hospital being provided in people’s own homes or place of residence instead [7] [8], with the “virtual ward” term often used for programmes that rely more heavily on technology. In April 2022, NHS England launched a national virtual wards programme with the aim of expanding the ability to provide acute (up to 14-days) hospital-level care at home (or normal place of residence) to patients who would otherwise be in hospital [9]. Providing either “step-up” care (enabling people to stay at home who might instead have been admitted) or “step-down” (enabling discharge of those who would otherwise have remained in hospital), the ambition was that all virtual wards would be technology-enabled [10]. The use of technology for remote monitoring was intended to enable patients (or their carers) to measure vital signs (e.g. through wearable technology). These readings are then transmitted to healthcare professionals for remote review. The NHS England virtual wards programme made £450 million of funding available over two years, and required that systems would “fully exploit remote monitoring technology and wider digital platforms to deliver effective and efficient care” [9]. Other UK nations have introduced similar schemes [11] [12] [13].

Since the introduction of the NHS England programme, virtual wards have expanded rapidly: between May 2022 and January 2023, the number of ‘beds’ available on virtual wards in England increased by 60% [14]. Many of the beds are on “frailty” virtual wards, which are intended to care for people aged over 65 with frailty-related illness. However, the adoption of remote monitoring technology in virtual wards caring for people with frailty (39% of wards) has been much lower than for other medical conditions [15], e.g. respiratory (67%) and circulatory (71%) [15]. The reasons for the relatively low uptake of remote monitoring in virtual wards caring for people with frailty are poorly understood. We aimed to address this deficit by exploring stakeholders’ views and experiences of implementing remote monitoring in frailty virtual wards.

## Methods

### Sampling and recruitment

Participants eligible to take part in interviews included those involved in or facilitating patient care in frailty virtual wards, and those engaged in policy, planning, managing, organising, or evaluating UK based frailty virtual wards. Participants were recruited to a study seeking to build a theory of change for frailty virtual wards from February 2024 to July 2024 through multiple channels including social media, an NHS England e-newsletter, and emails to professional networks supplemented by snowball sampling. We preferentially recruited participants with experience of working in England, but did not exclude those in other parts of the UK. This approach aimed to gather a diverse range of experiences and insights from different contexts.

### Data collection and analysis

Interviews and qualitative analysis were conducted by two experienced qualitative researchers using a semi-structured topic guide to explore stakeholders’ views and experiences of frailty virtual wards, including the use of remote monitoring technology. The average interview length was 40 minutes (range 23 -79 minutes). Interviews were digitally recorded, transcribed verbatim and anonymised. Qualitative analysis was based on the constant comparative method [16], supported by NVIVO. Initial open coding was used on a sub-selection of transcripts representing a range of different types of stakeholders. The coding framework was iteratively updated throughout the analytic process, with regular researcher debriefs held to identify opportunities for adjusting the interview topic guide and to discuss codes and themes. Recruitment continued until theoretical saturation was reached [17].

### Patient and public involvement

We recruited a patient and public involvement (PPI) group comprising six people either living with frailty or caring for someone living with frailty, running three workshops where participants provided feedback on what they felt was important to ensure frailty virtual wards worked for patients and carers and on considerations in designing and implementing remote monitoring.

### Ethical approval

Ethical approval was granted by the Cambridge University Psychology Research Ethics Committee (Application No: PRE.2023.128).

## Results

Fifty-five individuals expressed interest in taking part in interviews. Of these, it proved possible to complete interviews with 42 who met our eligibility criteria. Participants included 25 frontline healthcare staff working on frailty virtual wards, 7 researchers/evaluators, 5 NHS staff with a regional or local operational role in relation to frailty virtual wards, three individuals working at policy level, and two technology supplier/advisors for virtual wards. Table one provides demographic details of the participants interviewed in this study.

We highlight four key challenges associated with implementing remote monitoring into frailty virtual wards as identified in the 2022 NHS England national guidance: professional ambivalence about the suitability of remote monitoring for this patient group; the nature and extent of change in clinical practices and work system design; procuring the right tools and technology; and tensions between standardisation and local flexibility.

### Professional ambivalence about the role of remote monitoring in virtual wards caring for people with frailty

While 2022 NHS England guidance described remote monitoring as enabling patients to record their vital signs and sharing data via a remote platform, healthcare professionals in our study were largely in consensus that this type of technology was of limited value on virtual wards caring for people with frailty. Instead, professionals tended to prioritise clinical judgement and discussions with patients and carers over technology-derived data, and perceived multiple practical barriers to deploying remote monitoring for a frail population. In practice, remote monitoring was either not used or rarely used by most professionals we interviewed.

> *“We have not yet gone down the road of technology-enabled care, so we’re not doing remote monitoring. For lots of reasons, partly because we haven’t quite had the time and headspace to roll it out. But partly because I’m not sure of the value of numbers in managing frailty, it’s much more around judgement*.*” (participant 26, frontline healthcare professional)*

Among the practical barriers, participants identified that much of the available equipment for remote monitoring was unsuitable for people living with frailty, and that patients and their carers were often unable to collect and report data owing to cognitive or physical limitations, and that digital affordances in people’s homes were limited.

“[They] *don’t often have Wi-Fi at home, they’ve barely got a landline*.*” (participant 12, frontline healthcare professional)*

> *“We’ve piloted some wearables they call them, basically smartwatches, but they don’t seem to work on very frail skin and some of them are just physically quite heavy for people” (participant 42, frontline healthcare professional)*

Participants emphasised that the clinical complexity of people on frailty virtual wards meant that they were not appropriate candidates for remote monitoring and that a pragmatic and personalised approach was more appropriate.

> *“So, I think there’s something about being pragmatic about monitoring when it’s indicated, not monitoring for monitoring’s sake. And I think that’s something that NHSE in their spec and expectations, hasn’t understood for frailty, that they’ve set a target for how many patients are under technology-enabled care. And I don’t think that’s appropriate, certainly not from our lived experience of it*.*”* (participant 26, frontline healthcare professional)

### Adapting practices, responsibilities and roles for remote monitoring implementation

Participants identified that remote monitoring would require multiple adaptations of practices, responsibilities and roles on the part of patients, carers and staff. For example, patients or their informal carers would need to take a more proactive role in monitoring and managing their own health, with all the burdens that they would involve. Formal carers in patients’ homes were also reported to be reluctant to support remote monitoring, in part because of time pressures and in part because it involved new risks and responsibilities.

> *“It’s a complete change in the way of thinking, I think, isn’t it?” (participant 25, operational role at a regional/local level)*
>
> *“We met with their, the leads of them [paid carers] and it’s very much they’re up against it, they are very minutely timed for the tasks they have to carry out already and this [remote monitoring] was another thing… all reality with someone who’s frail with multiple layers of clothes. You might have to do it a few times over. It is a lengthier process. (participant 22, frontline healthcare professional)*

Healthcare professionals reported that remote monitoring had multiple implications for their own roles: they had to learn how to use the equipment themselves and had to be able to interpret and respond to the outputs from remote monitoring effectively, as well as educating patients/carers on how to use the systems. These overheads compounded the sense for many that remote monitoring for people with frailty in hospital-at-home settings was of limited value and involved many hidden resource costs.

> *“if you’re only going to be on the caseload for a day or two, by the time you’ve set it up and they’ve worked out how to use it, then you’re practically taking it down again*.*” (participant 20, frontline healthcare professional)*

> *“I suppose, what we’ve seen is sometimes patients, you know, being given the tech and haven’t even explained how to use it. They just get a bit anxious and they need that additional visit the next day to really be shown how to use it*.*” (participant 2, evaluator/researcher)*

The infrastructures and systems needed for remote monitoring were also perceived as under-developed. Where the team responsible for overseeing remote monitoring was separate from the team providing in-person care, for example, risks of missing important information or duplicating tasks were identified.

> *“they weren’t on our community care record to start with, so we couldn’t see what they were advising patients. And then again that risk of doubling up or missing out on something was too high*.*” (participant 28, frontline healthcare professional)*

Further challenges were associated with coordinating roles and responsibilities related to remote monitoring outside of core hours for virtual wards. Understandings and expectations among staff about how remote monitoring should be managed overnight and of the governance and risk management systems were reported to be often unclear or conflicting.

> *“…what happens if somebody is being monitored for 24 hours and something goes wrong in the middle of the night? We’re not around. So, there’s some governance issues around that, around relying on that monitoring when we’re not actually providing a service out of hours*.*” (participant 40, operational role at regional/local level)*

> “*We thought, oh, we wouldn’t be measuring all these things overnight in hospital unless they were on high dependency or intensive care and they’re not on high dependency and intensive care, so why do we need to do them*” *(participant 42, frontline healthcare professional)*

### Procuring the right tools and technology

A key challenge was seen to lie in ensuring that equipment and systems for remote monitoring were suitable for the clinical needs. While those at national level were clear about the need for procurement to be done right, in practice a multiplicity of suppliers, variable quality, and lack of interoperability were persistent challenges.

> *“there’s a baffling array of… system providers have really leapt into this space… But actually, not everything functions that effectively*.*” (participant 4, policy-maker)*

Because procurement of equipment was done largely locally, and generally without regard to larger-scale considerations, questions of how clinical information could be exchanged between other systems appeared to be unaddressed.

> *“I can only… I think the difficulty, in just speaking to lots of other people, is that primary care and secondary care don’t really speak to each other. So, it makes it quite tricky*.*With virtual wards, we’re the middle person. So, we’re not primary care, but they’re not… You know, they’re not in the hospital, in secondary care. So, it’s this middleman and making sure that… And I don’t think the technology is there for the communication between both*.*” (participant 39, frontline healthcare professional)*

Participants suggested that technology providers needed to innovate and adapt their products to meet the needs of patients with frailty and frailty virtual ward services, but that procurement processes and short-term funding cycles got in the way, forcing competition primarily on the basis of price and inhibiting partnerships between technology providers and frailty virtual ward services.

> “*One of the biggest barriers is the annual funding cycle. So if trusts only get a year’s funding, then they can’t have a ten year partnership with somebody…it makes it very precarious, particularly for the small, more innovative suppliers I think*.*” (participant 3, technology advisor)*

> *“So, the procurement department takes over, they just select someone, mainly based on price, I think it was more than 60% of the old tender was on price… what’s happening now is that the nurses and the clinicians and the doctors are really angry and say, well we don’t want to work with that party because we have invested a lot in that supplier*.*” (participant 18, technology supplier)*

### The tensions between standardisation and local flexibility

A further challenge in implementing remote monitoring in virtual wards caring for people with frailty lay in tensions between standardisation and flexibility. Evaluators of virtual wards highlighted that the heterogeneity of virtual wards, including variations in how remote monitoring was implemented, made it challenging to evaluate frailty virtual wards.

> *“*..*so there is an inherent challenge that we talk about frailty virtual wards as if that is a very well defined and very consistent thing regardless of where it is undertaken. And the reality is it’s not, these are very different services run in very different ways by very different criteria to admit and pathways*… *even though we’re referring to them as one group of services that we talk about as if they’re all the same*.*” (participant 14, researcher/evaluator)*

However, frontline healthcare professionals felt that efforts to standardise some elements, such as the requirement for services to use remote monitoring technology, or targets for the number of patients that should be using it, were often unhelpful.

> *“Initially we didn’t want any digital devices because we thought that patients wouldn’t use them. We had them because you have to have that in order to secure the funding, but as I said most patients don’t use them. We just ring up and we speak to them and that’s often the most beneficial part. So, I’d like to…well, I don’t want to say we were right, but we were right about that one. It’s the personal touch in the digital age that people appreciate that works best*.*” (participant 9, frontline healthcare professional)*

While policy-makers reported that they had initially enabled a flexible implementation approach, where services were able to design their own models of care and could select their own solutions, policy was now seeking to promote a more standardised model at scale based on best practice examples. Over time, for example, the view had emerged that a blend of face-to-face care alongside remote monitoring constituted good practice. Linked to this, the official terminology was evolving to shift from “virtual wards” to “Hospital at Home,” indicating less emphasis solely on technology.

> *“We’re now looking towards standardising that model and making sure that wherever you are across the country, you can access the same level of care. So, we would see that you would need a blended model with the ability to offer face-to-face care as well as that remote monitoring*.*” (participant 16, policy-maker)*
>
> *“We’re still transitioning, but in six months, we will not talk about virtual wards; we will only talk about Hospital at Home*.*” (participant 17, policy-maker)*

This move was broadly welcomed by the different participant groups, who felt that the term “virtual wards” was misleading.

> *“I also think that Hospital at Home is a more useful term, because virtual wards give the impression that people are literally just being monitored by robots, or certainly monitored from afar. While, in practice, a lot of the virtual wards have quite a high face to face ratio*.*” (participant 7, researcher/evaluator)*.

## Discussion

Current projections suggest that an additional 23,000 to 39,000 hospital beds will be required in England by 2030/31 [18]. Providing hospital care in people’s usual place of residence offers a promising alternative to building more NHS estate and potentially reduce some risks and improve efficiency [19]. While remote monitoring through technology has been a key ambition of a major NHS virtual wards programme, this qualitative study of stakeholders’ views and experiences suggest a number of key challenges in delivering on this aspiration in the care of people with frailty.

One of these challenges was that “convincing people that the solution chosen is the right one” [20] was largely not achieved for professionals caring for people on frailty virtual wards. Frailty is defined in the research literature as ‘a clinically recognisable state of increased vulnerability resulting from aging-associated decline in reserve and function across multiple physiologic systems such that the ability to cope with everyday or acute stressors is comprised’ [21]. Most health professionals that we interviewed were of the view that remote monitoring was, for a wide range of clinical, practical and social reasons, not appropriate for most patients with frailty.

A further challenge was that the work that professionals and patients/carers needed to do for remote monitoring to become routine practice [22] had not been sufficiently well understood, designed, or supported, nor was remote monitoring perceived as offering a relative advantage

[23] that would justify change. Also lacking was the infrastructure needed to ensure that the work involved in remote monitoring was coordinated and effective [24], with clear systems and responsibilities for communication and action. As previous studies have found, remote monitoring may increase staff workload, perhaps in ways that are not readily visible [25]. Also problematic were issues regarding tools and technology. The equipment clinicians and patients were expected to use was often regarded as sub-optimal, linked to procurement practices and short-term funding cycles that also stifled innovation.

These findings suggest some ways in which future national programmes involving technology might be improved. One of these is through use of a structured framework (see example in Table two) that would prompt attention to the range of considerations that might be relevant. More broadly, there is need to identify the models, resources and processes that are required, including work system design. While standardisation is increasingly seen as providing part of the solution to ensuring best practice in virtual wards [26], previous studies suggest that standardisation is likely to be much more difficult to achieve once practices and models have already become established [27]. Furthermore, it is important to distinguish between standardisation efforts which stifle innovation or lead to unintended consequences (i.e. “gaming” targets), and those which promote better patient outcomes and consistent quality of care [28]. As suggested in previous research, successful standardisation of virtual wards relies on policy makers achieving an optimal balance between standardisation and local flexibility [29].

Further recommendations centre on enabling services to adopt a cyclical process of planning, implementation, evaluation, and adaptation [30]. Essential to this process is co-designing solutions with patients, carers, and staff who care for patients with frailty across different health and social care sectors. Services need to avoid becoming locked into increasingly expensive contracts with technology suppliers, while also avoiding pricing structures that prevent investment in innovation. Training is also important, and strategies such as training staff together (i.e. all staff teams involved in facilitating or directly caring for patients on virtual wards), and using simulations with post simulation debriefs, can help clarify the roles and responsibilities of different stakeholders [31].

A strength of our study is the in-depth exploration of stakeholders’ perceptions of remote monitoring in frailty virtual wards. Furthermore, collating data from a range of stakeholders enabled the study to present a diverse range of perspectives. Although the study did not include patients and carers as interview participants, the use of a PPI group provided us with valuable insights that facilitated the development of the theory of change for remote monitoring (Figure one) and will facilitate further research with this group to understand patients and carers’ experiences of remote monitoring. However, the study also has limitations. It is plausible that the methods of recruitment may have biased the sample in favour of particular views, for example participants recruited and interviewed via online methods may view technology more favourably than participants recruited and interviewed using non-digital methods. In addition, while our study recruited participants from across the UK with the aim of exploring the views of individuals across diverse contexts, individuals outside of England and individuals in the North East of England were underrepresented. Although our analysis did not indicate that the views and experiences of participants greatly diverged depending on the region or country the participant was based in, a more geographically diverse sample may have highlighted important differences in perspectives across the UK.

## Conclusion

Implementing remote monitoring in frailty virtual wards is complex. Success depends on a thorough understanding of when remote monitoring is appropriate for frail patients, the establishment of clear protocols and workflows that enhance both patient outcomes and operational efficiency, and further research and development to identify best practices for remote monitoring.

**Table 1:**
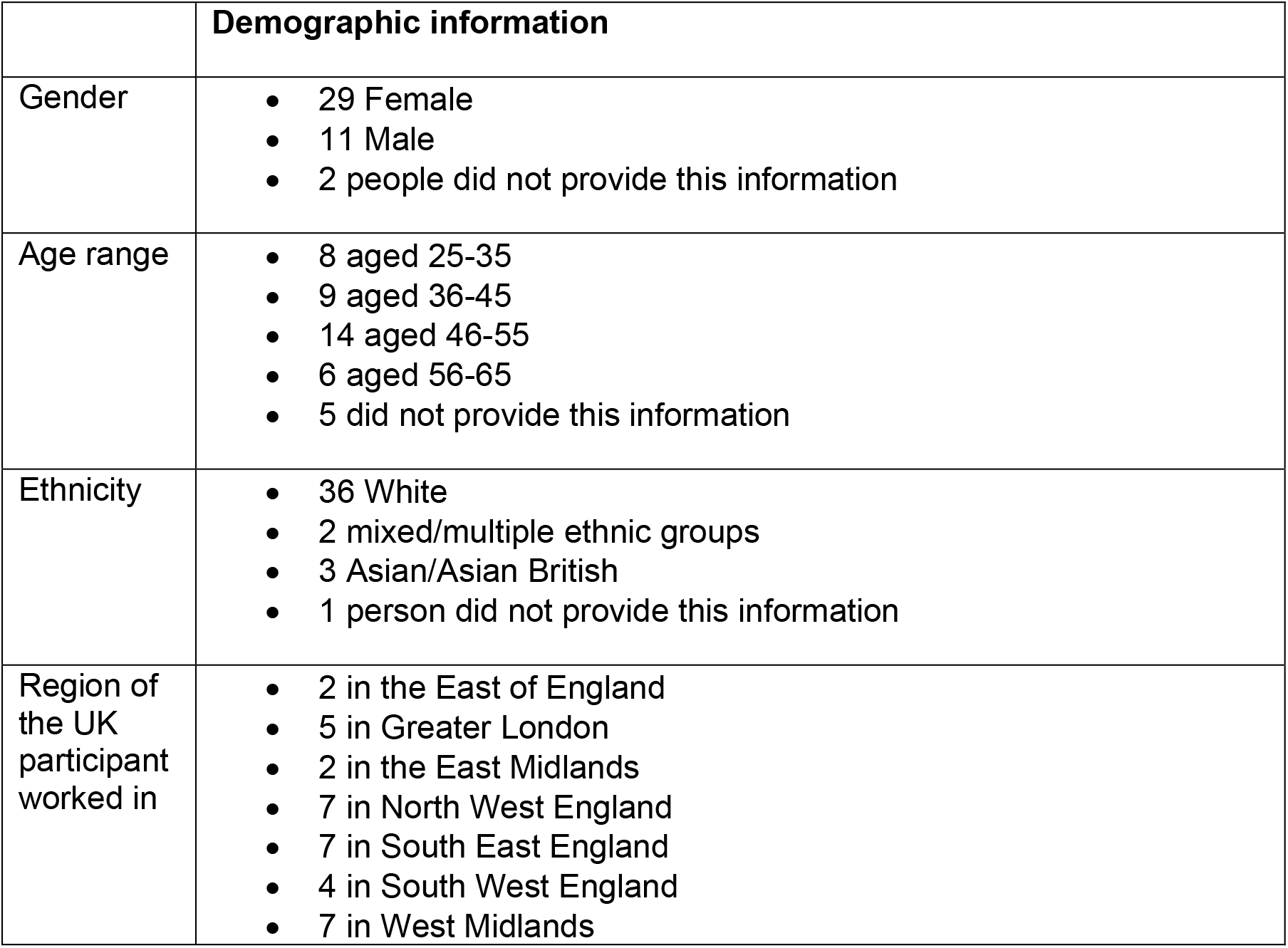
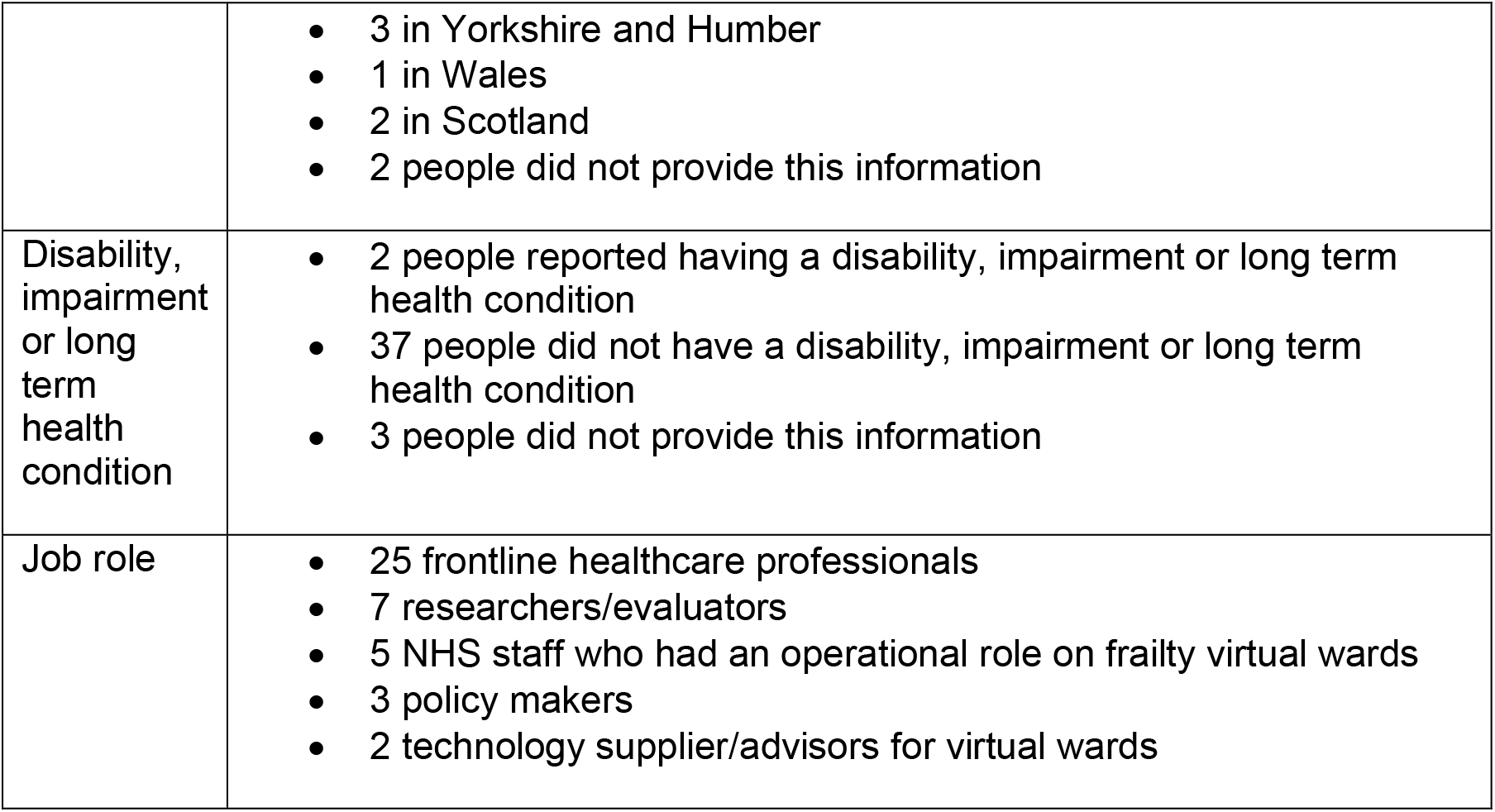
Participants’ demographic information.

**Table 2:**
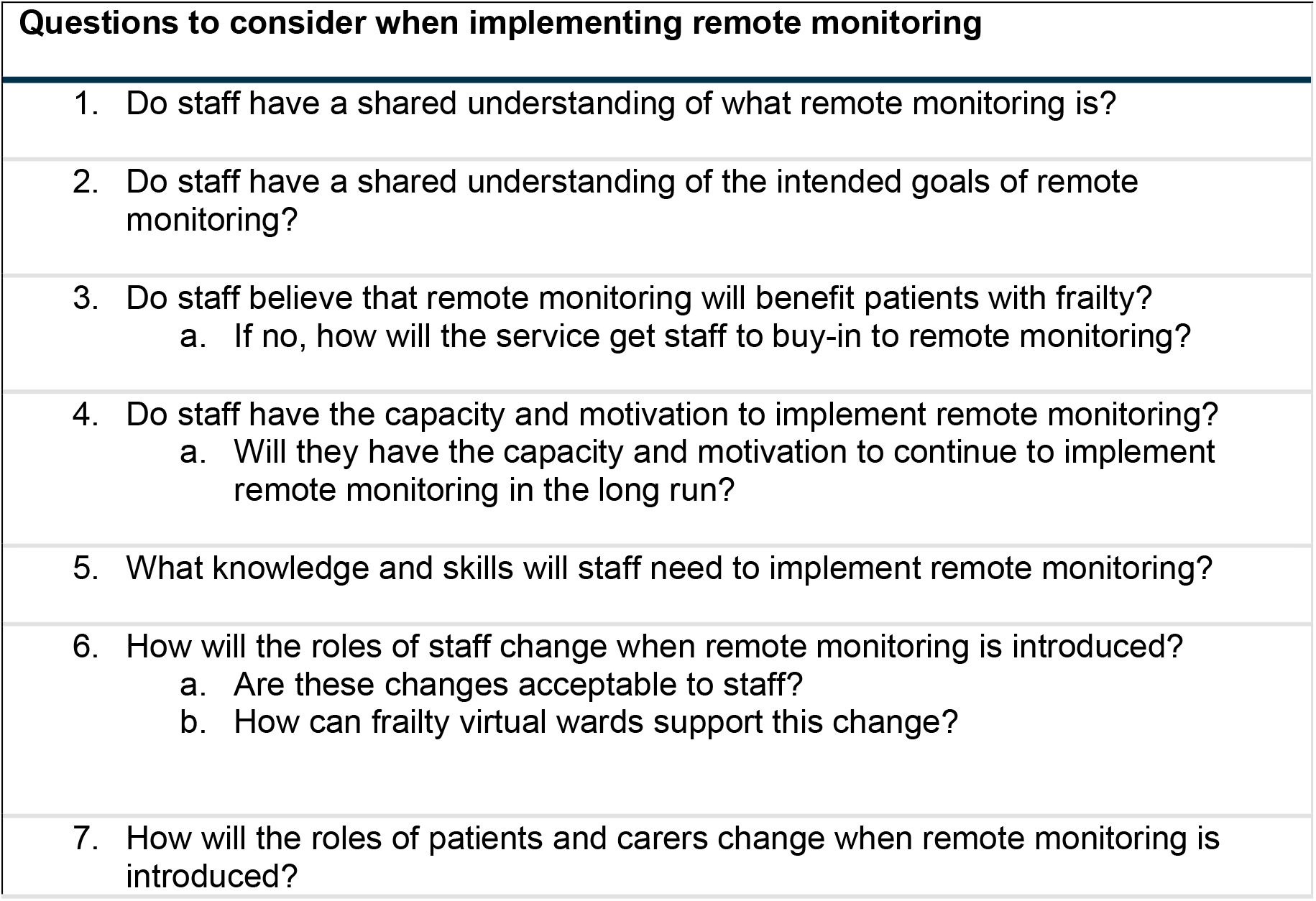
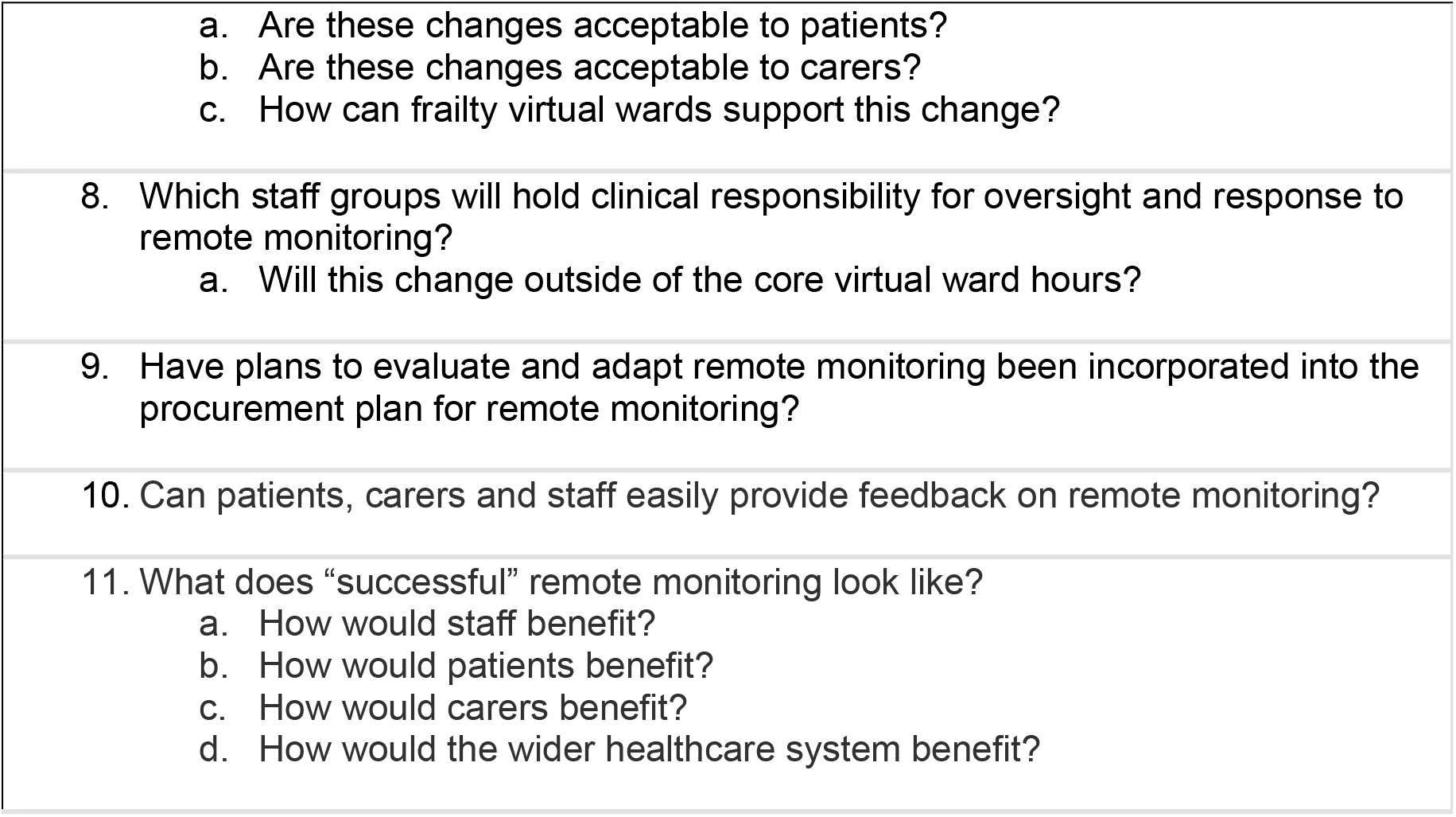
Questions to consider when implementing remote monitoring in frailty virtual wards.

| **Questions to consider when implementing remote monitoring** |  |
| --- | --- |
| 1. | Do staff have a shared understanding of what remote monitoring is? |
| 2. | Do staff have a shared understanding of the intended goals of remote monitoring? |
| 3. | Do staff believe that remote monitoring will benefit patients with frailty?<br>a. If no, how will the service get staff to buy-in to remote monitoring? |
| 4. | Do staff have the capacity and motivation to implement remote monitoring?<br>a. Will they have the capacity and motivation to continue to implement remote monitoring in the long run? |
| 5. | What knowledge and skills will staff need to implement remote monitoring? |
| 6. | How will the roles of staff change when remote monitoring is introduced?<br>a. Are these changes acceptable to staff?<br>b. How can frailty virtual wards support this change? |
| 7. | How will the roles of patients and carers change when remote monitoring is introduced? |

|  |
| --- |
| a. Are these changes acceptable to patients?<br>b. Are these changes acceptable to carers?<br>c. How can frailty virtual wards support this change? |
| 8. Which staff groups will hold clinical responsibility for oversight and response to remote monitoring?<br>a. Will this change outside of the core virtual ward hours? |
| 9. Have plans to evaluate and adapt remote monitoring been incorporated into the procurement plan for remote monitoring? |
| 10. Can patients, carers and staff easily provide feedback on remote monitoring? |
| 11. What does “successful” remote monitoring look like?<br>a. How would staff benefit?<br>b. How would patients benefit?<br>c. How would carers benefit?<br>d. How would the wider healthcare system benefit? |

## Data Availability

The data are not publicly available as sharing this data could potentially compromise participant privacy. For any further inquiries, please contact the corresponding author.

## Acknowledgments

We thank the study participants and members of the patient and public involvement group for their time and insights. We would also like to thank Wezzie Banda, for supporting the management of this project.

## References

1. Levi B, Borow M, Wapner L, Feldman Z. Home hospitalization worldwide and in Israel. Isr Med Assoc J 2019; 21(8): 565–7.

2. Royal College of Emergency Medicine. NHS crisis continues as hospital bed numbers near capacity. Royal College of Emergency Medicine 2024. Available from: https://rcem.ac.uk/nhs-crisis-continues-as-hospital-bed-numbers-near-capacity/ (23 September 2024, last accessed).

3. British Medical Association. NHS hospital beds data analysis. British Medical Association 2022. Available from: https://www.bma.org.uk/advice-and-support/nhs-delivery-and-workforce/pressures/nhs-hospital-beds-data-analysis (23 September 2024, last accessed).

4. Guest JF, Keating T, Gould D, Wigglesworth N. Modelling the annual NHS costs and outcomes attributable to healthcare-associated infections in England. BMJ Open 2020; 10(1): e033367.

5. Hadden C, Tse J. Corridor care: unsafe, undignified, unacceptable. Royal College of Nursing 2024.

6. Loyd C, Markland AD, Zhang Y, et al. Prevalence of hospital-associated disability in older adults: a meta-analysis. J Am Med Dir Assoc 2020 Apr; 21(4): 455–61.e5.

7. Hospital at Home Society. What is Hospital at Home? Hospital at Home Society 2023. Available from: https://www.hospitalathome.org.uk/whatis (23 September 2024, last accessed).

8. NHS. Supporting information: virtual ward including hospital at home. NHS 2022.

9. NHS. 2022/23 priorities and operational planning guidance. NHS 2022; 22 February 2022.

10. NHS. A guide to setting up technology-enabled virtual wards. NHS 2021. Available from: https://transform.england.nhs.uk/key-tools-and-info/a-guide-to-setting-up-technology-enabled-virtual-wards/ (4 July 2024, last accessed).

11. Scottish Government. Hospital at home for older people. Scottish Government 2024. Available from: https://www.gov.scot/news/hospital-at-home-funding/#:~:text=Hospital%20at%20Home%20enables%20people,consultant%20in%20their%20own%20home (23 September 2024, last accessed).

12. Welsh Government. Funding to increase allied health professionals and access to community-based care. Welsh Government 2023.

13. Colligan J. An economic assessment of the South Eastern Trust virtual ward. South Eastern Health and Social Care Trust (SEHSCT) 2015. Available from: https://www.rcn.org.uk/-/media/Royal-College-Of-Nursing/Documents/Professional-Development/Research/Innovations/Burdetters-Case-Studies/Janice-Colligan-Case-Study.pdf (23 September 2024, last accessed).

14. NHS England. World-leading NHS virtual wards treat 100,000 patients in a year. NHS England 2023. Available from: https://www.england.nhs.uk/2023/03/world-leading-nhs-virtual-wards-treat-100000-patients-in-a-year/ (23 September 2024, last accessed).

15. Chappell P, Co M, Hardie T, et al. What do virtual wards look like in England? The Health Foundation 2024.

16. Charmaz K. Constructing grounded theory: a practical guide through qualitative analysis. London: Sage, 2006.

17. Glaser BG, Strauss AL. The discovery of grounded theory: strategies for qualitative research. Chicago: Aldine Transaction, 1967.

18. Rocks S, Rachet-Jacquet L. How many hospital beds will the NHS need over the coming decade? The Health Foundation 2022.

19. Serrano LP, Maita KC, Avila FR, et al. Benefits and challenges of remote patient monitoring as perceived by health care practitioners: a systematic review. Perm J 2023; 27(4): 100–11.

20. Dixon-Woods M, McNicol S, Martin G. Ten challenges in improving quality in healthcare: lessons from the Health Foundation’s programme evaluations and relevant literature. BMJ Qual Saf 2012; 21(10): 876–84.

21. Xue Q-L. The frailty syndrome: definition and natural history. Clin Geriatr Med 2011; 27(1): 1–15.

22. Murray E, Treweek S, Pope C, et al. Normalisation process theory: a framework for developing, evaluating and implementing complex interventions. BMC Med 2010; 8(1): 63.

23. Leonardi P. When flexible routines meet flexible technologies: affordance, constraint, and the imbrication of human and material agencies. MIS Q 2011; 35(1): 147–67.

24. O’Reilly P, Lee SH, O’Sullivan M, et al. Assessing the facilitators and barriers of interdisciplinary team working in primary care using normalisation process theory: an integrative review. PLoS One 2017; 12(5): e0177026.

25. Davis MM, Freeman M, Kaye J, et al. A systematic review of clinician and staff views on the acceptability of incorporating remote monitoring technology into primary care. Telemed J E Health 2014 May; 20(5): 428–38.

26. NHS England. Virtual wards operational framework. NHS England 2024. Available from: https://www.england.nhs.uk/long-read/virtual-wards-operational-framework/ (23 September 2024, last accessed).

27. Kriznik NM, Lamé G, Dixon-Woods M. Challenges in making standardisation work in healthcare: lessons from a qualitative interview study of a line-labelling policy in a UK region. BMJ Open 2019; 9(11): e031771.

28. Best A, Greenhalgh T, Lewis S, et al. Large-system transformation in health care: a realist review. Milbank Q 2012; 90(3): 421–56.

29. Hakim R. Realising the potential of virtual wards. London: NHS Confederation 2023.

30. NHS. First steps towards quality improvement: a simple guide to improving services. NHS Improving Quality 2014.

31. Weller J, Boyd M, Cumin D. Teams, tribes and patient safety: overcoming barriers to effective teamwork in healthcare. Postgrad Med J 2014; 90(1061): 149–54.

